# Watching Over Life: Practices and Meanings of Postpartum Vital Signs Monitoring in Eastern Uganda

**DOI:** 10.1101/2025.05.13.25327548

**Authors:** Jonathan Babuya, Frank Kayemba, Sinani Waiswa, Allan G. Nsubuga, Helen Ewing, Irene Atuhairwe, Esther Ijangolet, Eric Otim, Kibuuka Ronald, Atugonza Jesca, Francis Okello, Prossy Nakattudde, Faith Nyangoma, Betty Nakawuka, Nichola Kabahinda, Assen Kamwesigye, Jill Apoya, Paul Waako, Julius Wandabwa, Milton Musaba, Ahaisibwe Bonaventure, Tod Aeby, Sarah Nandutu, Kenneth Mugabe, Kennedy Pangholi, Doreck Nahurira, Brian Agaba, Immaculate Mbwali, Johnson Bahati, Enid Kawala Kagoya

**Author notes:** **Corresponding author** (J.B).

## Abstract

**Introduction:** Postnatal deaths account for about one-third of maternal mortality in Low- and middle-income countries (LMICs). Nearly half occur within the first 24 hours and are preventable through continuous inpatient monitoring. This study assessed maternal mortality within the first 24 postnatal hours; and the current postpartum monitoring practices, including the availability of monitoring charts and the frequency of vital sign assessments in eastern Uganda.

**Methods:** This cross-sectional study reviewed the medical records of all postpartum women admitted at Mbale Regional Referral Hospital, Eastern Uganda, between August 2022 and February 2023. A semi-structured data extraction form was used. The outcome variable was postpartum monitoring; categorized as “No” for women with no vital signs monitored and “Yes” for those with at least one vital sign monitored. Descriptive analysis was performed using Stata Version 17.0, and Poisson regression was employed to explore the association between various factors and postpartum monitoring.

**Results:** Medical records of 2,717 postpartum mothers were reviewed. The median (interquartile range) age was 25(20-30) years. A minority (34%; 910/2717) of mothers had vital sign observation charts in their medical records and very few (4%; 110/2717) had a fluid balance chart. About 24% (651/2717) of mothers had at least one vital sign recorded. Blood pressure was the most (16.8%; 456/2717) recorded vital sign followed by pulse rate (13.6%; 369/2717). Respiratory rate and temperature were the least monitored vital signs with 1.2% and 1.1% records respectively. Mothers who delivered by C-section had a 151% higher prevalence of postpartum monitoring compared to those who had a vaginal delivery (aPR: 2.51; 95% CI: 2.19–2.89). Maternal mortality rate on the first postpartum day was 8.8 per 1000, with postpartum hemorrhage accounting for 61%.

**Conclusion:** Postpartum vital signs monitoring was low. LMICs need to devise means of automating the monitoring process.

## Introduction

Postpartum care guidelines are designed to promote early detection and management of common puerperal complications, consequently reducing maternal and newborn mortality and morbidity, and providing a positive postnatal experience [1]. Despite the universal presence of these guidelines, maternal mortality remains disproportionately high, with Low- and middle-income countries (LMICs) accounting for 95% of global deaths from preventable conditions related to pregnancy and childbirth [2-4]. For instance, the lifetime risk of maternal death in sub-Saharan Africa is 1 in 40, compared to 1 in 16,000 in Australia or 1 in 65,000 in Belarus [4]. Inadequate postpartum care is a key contributor to this mortality. In sub-Saharan Africa, about one-third of maternal deaths occur during postpartum, with rates as high as 60% in some countries [5].

Approximately 50% of the postnatal maternal deaths occur within the first 24 hours, with postpartum hemorrhage (PPH) being the leading cause, followed by embolic events and hypertensive disorders such as eclampsia [6]. To address this, World Health Organization (WHO) postpartum care guidelines recommend continuous in-patient monitoring for the first 24 hours postpartum for; vaginal bleeding, uterine tone, fundal height, temperature, blood pressure, heart rate (pulse), and urine output [1]. These parameters aid early detection of postpartum complications. Recording and interpreting them was eased by developing standardized tools, and indices like the Early Obstetric Warning Score (EOWS), Obstetric Shock Index (OSI), and Fluid Balance Chart [7-11]. Using these tools significantly improves detection of postpartum complications like PPH [8, 9, 12, 13]. Though continuous monitoring of postpartum mothers is efficacious, its implementation in LMICs is questionable. Most LMICs experience a shortage of healthcare workforce and monitoring devices among other challenges, which may hinder them from providing the recommended postnatal care package [14-16]. Given these challenges, there is a need to assess the current postnatal monitoring practices in LMICs and their alignment with established guidelines.

In this study, we assessed for 1) the availability of vital sign monitoring charts and fluid balance charts at the postnatal ward of a tertiary health facility in eastern Uganda; 2) the frequency of monitoring of vital signs among postpartum women in the first 24 hours post-delivery; 3) the causes of maternal death in first 24 hours postpartum. The national guidelines in Uganda recommend vital signs monitoring every 30 minutes in the first 2 hours postpartum, and 6-hourly for the following 24 hours. They also recommends hourly monitoring for bleeding for the first 6 hours [17].

## Methods

### Study Design

This was a cross-sectional study. Medical records of all postpartum women admitted at postnatal ward at Mbale Regional Referral Hospital (Mbale RRH) from August 2022 to February 2023 were reviewed.

### Study area

The study was conducted at Mbale Regional Referral Hospital, which is located at the Centre of Mbale city, Elgon zone in eastern Uganda. It is a tertiary care and the highest-level referral facility in the region offering both general and specialized healthcare services in out-patient and inpatient departments. Mbale RRH is a government/ public hospital with a 450-bed capacity, 360 staff, and a catchment area of 16 districts with a population of 4.6 million. Over 12,000 deliveries are conducted annually [18]. The hospital uses hand-written, paper-based medical records to store patient data.

### Study population

All postpartum mothers who were admitted to the postnatal ward at Mbale RRH.

### Sample size calculation

We reviewed medical records of all post-partum mothers admitted at Mbale RRH in 6 months from August 2022 to February 2023. Charts of 2,717 mothers were reviewed. We choose 6 months to extract a sufficient and representative sample of cases within the hospital’s routine operations.

### Inclusion criteria

The study included medical records of all postpartum mothers admitted at Mbale RRH from August 2022 to February 2023.

### Exclusion criteria

Medical records with evidence of lost records such as plucked-off paper.

### Sampling criteria

All medical records of postpartum mothers admitted to the postnatal ward from August 2022 to February 2023 were included.

### Data collection tool

We developed a data extraction form based on the postnatal monitoring recommended by the Uganda postpartum care guidelines [17]. The form had three sections; the first section recorded the mother’s biodata, mode of delivery and outcome of delivery; the second section assessed the presence of vital sign observation and fluid balance charts, and frequency of monitoring for vitals as per the guidelines: respiratory rate, temperature, blood pressure, pulse rate, oxygen saturation, urine output and blood loss. The last section recorded the maternal outcomes at 24 hours. The data extraction form was validated using a sample of 24 mothers’ medical records to ensure completeness. The form was uploaded in an electronic data collection tool: KoBo Toolbox which is an open-source software developed by the Harvard Humanitarian Initiative with support from United Nations agencies, CISCO, and partners to support data management by researchers and humanitarian organizations (https://www.kobotoolbox.org/).

### Primary outcome

The outcome variable was postpartum monitoring, denoted as a “Yes” if a mother had at least one vital sign at least once monitored and as a “No” if no vital sign was monitored.

### Data collection methods

Data were extracted by 8 research assistants over two weeks. They were briefed on the study protocol. They were also pre-trained on using the electronic questionnaire and were tested for uniformity in data collection. A pilot test was conducted on 50 records to ensure consistency before actual data collection started. Data were reviewed to ensure complete filling.

### Data analysis

The dataset was cleaned in Stata Version 17.0. Descriptive statistics were used to summarize study population characteristics and clinical outcomes. Continuous variables were summarized using mean and standard deviation (SD), while categorical variables were summarized using frequencies and percentages (%). Graphical representation was done using R. Bivariate and Multivariate analyses were conducted using a generalized linear model for the Poisson family with a log link to assess the strength of association using prevalence ratios between selected factors and postpartum monitoring. Variables included in the multivariable model were based on clinical plausibility. Factors with a p-value less than 0.05 at multivariable regression analysis were considered significant.

### Ethical considerations

This study was conducted according to the Declaration of Helsinki and the principles of Good Clinical Practice and Human Subject Protection. We also sought permission from the hospital, maternity and postnatal ward in-charges, and the records offices of Mbale Regional Referral Hospital to access the participants’ charts. Patient-specific identifiers like Name and hospital ID were not captured.

Ethical approval was sought and obtained from the Research Ethics Committees (REC) of Mbale Regional Referral Hospital No. MRRH-2023-300. Written informed consent was obtained from all subjects or participants and the consent form was approved by Mbale Regional Referral Hospital Research and Ethics Committee (REC), MRRH 2023-300.

## Results

### Maternal demographics and clinical outcomes

Overall, medical records of 2,717 postpartum mothers were extracted. The median age of the mothers was 25 years, with an interquartile range of 20-30 years. Of the mothers, 19% (503/2717) were referred from lower-level facilities. Regarding the delivery method, 66% (1797/2717) of mothers had delivered vaginally, while 34% (920/2717) had had a caesarean section. Over 97% (2642/2717) of births had been live births, with 3% (75/2717) being stillbirths. The outcome variable was postpartum monitoring; 24% (651/2717) of mothers had at least one vital sign recorded Table 1. These are summarised in **Table 1**.

**Table 1:** Characteristics and clinical outcomes of postpartum mothers at Mbale Regional Referral Hospital, Eastern Uganda.

|  | **Postpartum monitoring:** At least one vital sign was recorded at least once. |  |  |
| --- | --- | --- | --- |
|  | No | Yes | Total |
|  | N=2066 (76%) | N=651 (24%) | N=2717 |
| **Age** |  |  |  |
| <20 | 368 (18%) | 104 (16.0%) | 472 (17.4%) |
| 20-29 | 1172 (56.7%) | 370 (56.8%) | 1542 (56.8%) |
| 30-39 | 488 (23.6%) | 162 (24.9%) | 650 (23.9%) |
| 40+ | 38 (1.8%) | 15 (2.3%) | 53 (2.0%) |
| **Religion** |  |  |  |
| Christian | 1118 (54.1%) | 320 (49.2%) | 1438 (52.9%) |
| Moslem | 555 (26.9%) | 164 (25.2%) | 719 (26.5%) |
| Not recorded | 378 (18.3%) | 163 (25.0%) | 541 (19.9%) |
| Others | 15 (0.7%) | 4 (0.6%) | 19 (0.7%) |
| **Parity** |  |  |  |
| Primiparas | 804 (38.9%) | 245 (37.6%) | 1049 (38.6%) |
| Para 2 to 4 | 972 (47.0%) | 306 (47.0%) | 1278 (47.0%) |
| Grand Parus | 267 (12.9%) | 96 (14.7%) | 363 (13.4%) |
| Great grand Parus | 23 (1.1%) | 4 (0.6%) | 27 (1.0%) |
| **Referral status** |  |  |  |
| No | 1760 (85.3%) | 445 (69.0%) | 2205 (81.4%) |
| Yes | 303 (14.7%) | 200 (31.0%) | 503 (18.6%) |
| **Mode of delivery** |  |  |  |
| Vaginal | 1529 (74.0%) | 268 (41.2%) | 1797 (66.1%) |
| C-section | 537 (26.0%) | 383 (58.8%) | 920 (33.9%) |
| **Foetal outcome** |  |  |  |
| Fresh Stillbirth | 16 (0.8%) | 23 (3.5%) | 39 (1.4%) |
| Live birth | 2025 (98.0%) | 617 (94.8%) | 2642 (97.2%) |
| Macerated Stillbirth | 25 (1.2%) | 11 (1.7%) | 36 (1.3%) |
| **Vital sign observation chart availability** |  |  |  |
| No | 1461 (70.7%) | 346 (53.1%) | 1807 (66.5%) |
| Yes | 605 (29.3%) | 305 (46.9%) | 910 (33.5%) |
| **Fluid balance chart availability** |  |  |  |
| No | 2034 (98.5%) | 573 (88.0%) | 2607 (96.0%) |
| Yes | 32 (1.5%) | 78 (12.0%) | 110 (4.0%) |
| **Mother's Outcome after 24 hours** |  |  |  |
| Died | 6 (0.3%) | 18 (2.8%) | 24 (0.9%) |
| Run away | 8 (0.4%) | 2 (0.3%) | 10 (0.4%) |
| improved (Discharged) | 2052 (99.3%) | 631 (96.9%) | 2683 (98.7%) |

### Availability of Vital sign observation chart

A minority (34%; 910/2717) of mothers had vital sign observation charts in their medical records, of these, only 9.3% (92/910) had been used/filled **(Table 1)**.

### Availability of Fluid balance chart

Very few (4%; 110/2717) mothers had a fluid balance chart in their records, of these, over 47% (60/110) had been used/filled **(Table 1)**.

### Frequency of monitoring for the different vital signs

About 24% (651/2717) of mothers had at least one vital sign recorded **(Table 1)**. Blood pressure was the most (16.8%; 456/2717) recorded vital sign followed by pulse rate (13.6%; 369/2717). Respiratory rate and temperature were the least monitored vital signs with 1.2% and 1.1% records respectively **(Table 2) (Fig 1) (Fig 2)**.

**Table 2:**
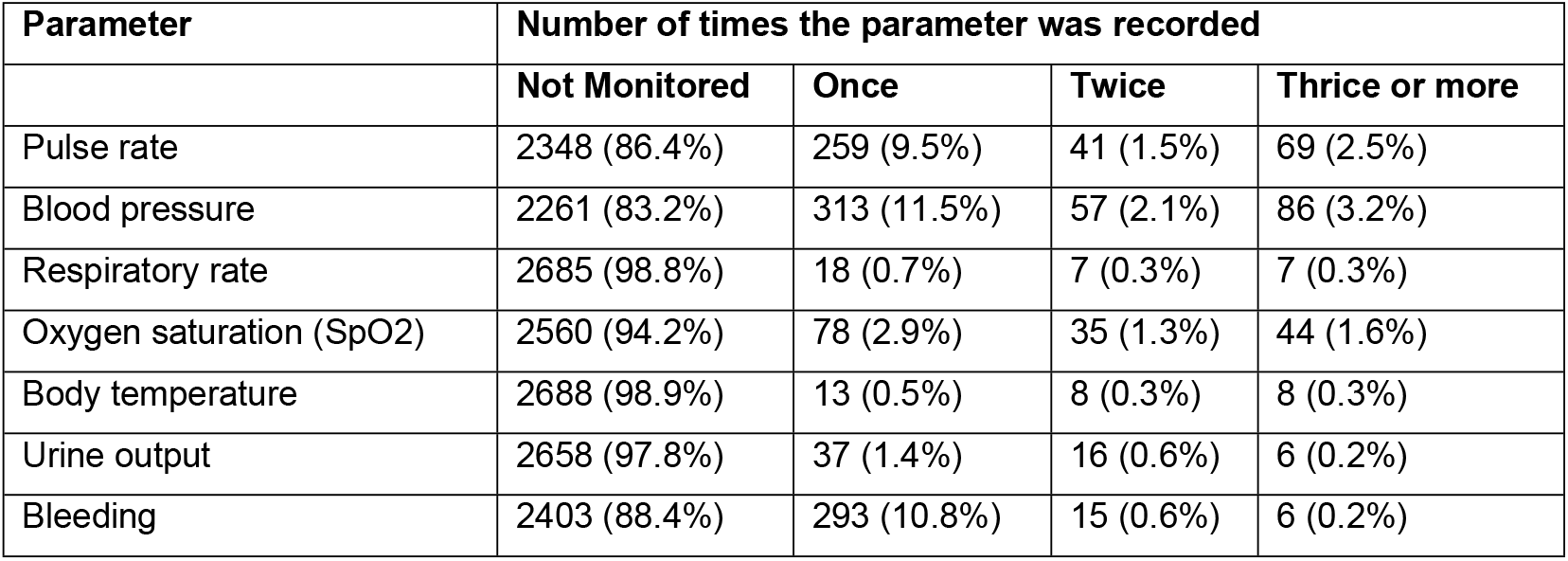
Frequency of Vital signs monitoring in the first 24 hours postpartum at Mbale Regional Referral Hospital, Eastern Uganda.

| Parameter | Number of times the parameter was recorded |  |  |  |
| --- | --- | --- | --- | --- |
|  | Not Monitored | Once | Twice | Thrice or more |
| Pulse rate | 2348 (86.4%) | 259 (9.5%) | 41 (1.5%) | 69 (2.5%) |
| Blood pressure | 2261 (83.2%) | 313 (11.5%) | 57 (2.1%) | 86 (3.2%) |
| Respiratory rate | 2685 (98.8%) | 18 (0.7%) | 7 (0.3%) | 7 (0.3%) |
| Oxygen saturation (SpO<sub>2</sub>) | 2560 (94.2%) | 78 (2.9%) | 35 (1.3%) | 44 (1.6%) |
| Body temperature | 2688 (98.9%) | 13 (0.5%) | 8 (0.3%) | 8 (0.3%) |
| Urine output | 2658 (97.8%) | 37 (1.4%) | 16 (0.6%) | 6 (0.2%) |
| Bleeding | 2403 (88.4%) | 293 (10.8%) | 15 (0.6%) | 6 (0.2%) |

**Fig 1.** Show the frequencies of vital sign monitoring in the first 24 hours postpartum at Mbale Regional Referral Hospital, Eastern Uganda.

**Fig 2.** Shows the frequencies of vital sign monitoring in the first 24 hours postpartum at Mbale Regional Referral Hospital, Eastern Uganda.

### Maternal mortality

Postnatal maternal mortality rate on the first postpartum day was 8.8 per 1000.

### Causes of maternal mortality

Causes of maternal death were recorded in 18 of the 24 deaths. They included Postpartum Hemorrhage and its complications 61% (11/18); Puerperal sepsis 11% (2/11); Pre-eclampsia 11% (2/11); Antepartum Hemorrhage 6% (1/11); Electrolyte imbalance 6% (1/11); cardiac arrest 6% (1/11). Summarized in **Fig 3**.

**Fig 3.** Shows the frequencies of vital sign monitoring in the first 24 hours postpartum at Mbale Regional Referral Hospital, Eastern Uganda.

### Factors associated with postpartum monitoring in the first 24 hours

Mothers who delivered by C-section had a 151% higher prevalence of being monitored in the first 24 hours postpartum compared to those who had a vaginal delivery (aPR: 2.51; 95% CI: 2.19–2.89). Availability of vital sign observation charts was associated with a 64% higher prevalence of vital signs monitoring (aPR: 1.64; 95% CI: 1.44–1.87). Availability of fluid balance chart was associated with a 54% higher prevalence of postpartum monitoring (aPR: 1.54; 95% CI: 1.29–1.83). Mothers referred from other facilities had a 63% higher prevalence of postpartum monitoring (aPR: 1.63; 95% CI: 1.42–1.87). Mothers with a fresh stillbirth (FSB) had a 51% higher prevalence of postpartum monitoring compared to those with a live birth (aPR: 1.51; 95% CI: 1.05–2.16). Summarized in **table 3**.

**Table 3:** Factors associated with Vital signs monitoring among postpartum mothers in the first 24 hours at Mbale Regional Referral Hospital, Eastern Uganda.

| Variable | cPR [95% CI] | P-value | aPR [95% CI] | P-value |
| --- | --- | --- | --- | --- |
| **Mode of Delivery** |  |  |  |  |
| Vaginal | 1 |  | 1 | 1 |
| C-section | **1.03[0.87-1.18]** | **<0.001** | **2.51[2.19-2.89]** | **<0.001** |
| **Outcome of Delivery** |  |  |  |  |
| **Live birth** | 1 |  | 1 |  |
| FSB | **0.93[0.51-1.34]** | **<0.001** | **1.51[1.05-2.16]** | **0.026** |
| MSB | 0.27[-0.33-0.86] | 0.38 | 1.41[0.86-2.32] | 0.179 |
| **Vital Signs Observation chart availability** |  |  |  |  |
| No | 1 |  | 1 |  |
| Yes | **0.56[0.41-0.71]** | **<0.001** | **1.64[1.44-1.87]** | **<0.001** |

| **Fluid Balance chart availability** |  |  |  |  |
| --- | --- | --- | --- | --- |
| No | 1 |  | 1 |  |
| Yes | **1.17[0.93-1.41]** | **<0.001** | **1.54[1.29-1.83]** | **<0.001** |
| **Referral Status** |  |  |  |  |
| No | 1 |  | 1 |  |
| Yes | **0.68[0.51-0.84]** | **<0.001** | **1.63[1.42-1.87]** | **<0.001** |

## Discussion

Our study shows gaps in the continuous monitoring of vital signs among postpartum mothers at Mbale Regional Referral Hospital. Despite established guidelines recommending up to 6 times as; monitoring for evidence of bleeding by inspecting the vulva half hourly for 2 hours, then 6-hourly for 24 hours; checking fundal height half hourly for two hours, and 6 hourly for 24 hours; observing and measuring vital signs including blood pressure, pulse, respiration rate and level of consciousness (half hourly for 2 hours, then every 6 hours for 24 hours) within the first 24 hours postpartum[19] to detect complications early, our findings indicate that only 24% of mothers had at least one recorded vital sign. This low rate of monitoring and documentation raises concerns about the quality of postnatal care and the ability of healthcare workers to promptly detect and manage postpartum complications.

Similar challenges in postpartum monitoring have been reported in other studies conducted in low-resource settings. For instance, Mugyenyi et al. (2021) assessed vital sign monitoring in a Ugandan regional referral hospital and found that the frequency of postpartum assessments was significantly lower than national and WHO recommendations, corroborating our findings[20]. Likewise, Kruk et al. (2016) analyzed maternal care functions in five African countries and found that basic maternal care, including vital sign monitoring, was often inconsistent, highlighting systemic challenges in postnatal care[14]. Beňová et al. (2023) also reported that inadequate postpartum monitoring is a major barrier to implementing effective maternal health interventions in sub-Saharan Africa[21]. This low rate of monitoring could be associated to multiple factors ranging from systemic shortages like low staffing, lack of equipment, low motivation and inadequate training of health workers, lack of stringent accountability and supervision for the health workers on duty and many more systemic factors that need to be investigated further to guide solutions and changes in the health sector to address this challenge.

Blood pressure (16.8%) and pulse rate (13.6%) were the most frequently recorded vital signs, while respiratory rate (1.2%) and temperature (1.1%) were the least monitored. These results align with those from other studies who observed that blood pressure was more commonly recorded due to its role in detecting hypertensive disorders, while other vital signs were frequently overlooked[22]. In contrast, Ibáñez-Lorente et al. (2021) found that in settings where a maternal early warning system was implemented, postpartum vital signs were monitored more consistently, leading to better maternal outcomes[7]. These findings suggest that structured monitoring protocols and digital tracking systems could improve adherence to postpartum surveillance guidelines.

Maternal mortality in the first 24 hours postpartum in our study was 8.8 per 1,000, with postpartum hemorrhage (PPH) accounting for 61% of the deaths. This finding is consistent with global trends, as WHO and Dol et al. (2022) identify PPH as the leading cause of maternal mortality worldwide, particularly within the first 24 hours postpartum [6]. However, studies from high-income settings report much lower postpartum mortality rates, attributed to timely interventions facilitated by continuous monitoring and digital alert systems [13]. The low rate of vital sign monitoring in our study suggests that many cases of hemorrhage, sepsis, and hypertensive disorders may not have been detected early enough for timely intervention.

Another key finding was that mothers who delivered by cesarean section (C-section) had a 151% higher prevalence of postpartum monitoring compared to those who had a vaginal delivery. This aligns with Agaba et al. (2024), who reported that postoperative patients receive closer monitoring due to the perceived risk of surgical complications [23]. However, this disparity suggests a need to strengthen routine monitoring for all postpartum women, regardless of delivery mode, as complications such as PPH and hypertensive disorders also occur after vaginal deliveries.

A major barrier to effective postpartum monitoring in our setting is the limited availability of structured monitoring tools. Only 34% of mothers had a vital sign observation chart, and just 4% had a fluid balance chart documented. This challenge has been widely reported in LMICs, where resource constraints and paper-based systems hinder systematic monitoring [7, 9]. Studies suggest that adopting digital solutions, such as electronic medical records with automated alerts, could improve adherence to monitoring protocols and enhance early detection of complications [9]. Additionally, structured training programs for healthcare workers on the use of Early Obstetric Warning Scores (EOWS) and Obstetric Shock Index (OSI) have been shown to improve early detection and management of postpartum complications [9, 13].

## Conclusion and Recommendations

Postpartum vital sign monitoring a crucial part of maternal care is still low in facilities, particularly among women who deliver vaginally. Strengthening adherence to WHO and national postpartum care guidelines is crucial. Interventions such as digital monitoring systems, staff training on postpartum surveillance, and increased availability of monitoring tools could improve maternal outcomes. Additionally, increasing the availability and utilization of vital sign observation charts and integrating digital tracking tools could help bridge the gap in postpartum care.

More studies should explore the feasibility of automated postpartum monitoring systems in low-resource settings, as well as the impact of healthcare worker training on maternal early warning systems in improving maternal outcomes.

## Data Availability

All relevant data are within the manuscript and its Supporting Information files.

## Declarations

All authors have nothing to declare.

## Acknowledgement

This study was supported by Seed Global Health in partnership with Mbale Regional Referral Hospital and Busitema University. We thank all the research team members for their valuable input and support.

## Supporting information

**S1 Dataset**

**S2 Figs. 1, 2 and 3**

**S3 Data collection tool**

## References

1. World Health Organization IC. WHO recommendations on maternal and newborn care for a positive postnatal experience. 2022.

2. WHO. WHO Factsheets: Maternal mortality 2024 [cited 2025 01/03/2025]. Available from: https://www.who.int/news-room/fact-sheets/detail/maternal-mortality.

3. Say L, Chou D, Gemmill A, Tunçalp Ö, Moller A-B, Daniels J, et al. Global causes of maternal death: a WHO systematic analysis. The Lancet Global Health. 2014;2(6):e323–e33. doi: 10.1016/S2214-109X(14)70227-X.

4. World Health Organization G. Trends in maternal mortality 2000 to 2020: estimates by WHO, UNICEF, UNFPA, World Bank Group and UNDESA/Population Division. 2023.

5. Merdad L, Ali MM. Timing of maternal death: Levels, trends, and ecological correlates using sibling data from 34 sub-Saharan African countries. PLoS One. 2018;13(1):e0189416. Epub 20180117. doi: 10.1371/journal.pone.0189416. PubMed PMID: 29342157; PubMed Central PMCID: PMCPMC5771557.

6. Dol J, Hughes B, Bonet M, Dorey R, Dorling J, Grant A, et al. Timing of maternal mortality and severe morbidity during the postpartum period: a systematic review. JBI Evid Synth. 2022;20(9):2119–94. Epub 20220901. doi: 10.11124/jbies-20-00578. PubMed PMID: 35916004; PubMed Central PMCID: PMCPMC9594153.

7. Ibáñez-Lorente C, Casans-Francés R, Bellas-Cotán S, Muñoz-Alameda LE. Implementation of a maternal early warning system during early postpartum. A prospective observational study. PLoS One. 2021;16(6):e0252446. Epub 20210603. doi: 10.1371/journal.pone.0252446. PubMed PMID: 34081737; PubMed Central PMCID: PMCPMC8174734.

8. Oyoo P, Qureshi Z, Osoti A, Gwako G, Mugambi JK, Okore J, et al. Blood loss monitoring chart, a game changer in postpartum hemorrhage detection and treatment: A direct observation study in 7 E-MOTIVE trial hospitals in Kenya. Journal of Obstetrics and Gynaecology of Eastern and Central Africa. 2024.

9. Nathan HL, El Ayadi A, Hezelgrave NL, Seed P, Butrick E, Miller S, et al. Shock index: an effective predictor of outcome in postpartum haemorrhage? Bjog. 2015;122(2):268–75. doi: 10.1111/1471-0528.13206. PubMed PMID: 25546050.

10. Agaba DC, Lugobe HM, Migisha R, Jjuuko M, Saturday P, Kisombo D, et al. Abnormal obstetric shock index and associated factors among immediate postpartum women following vaginal delivery at a tertiary hospital in southwestern Uganda. BMC Pregnancy and Childbirth. 2024;24(1):31. doi: 10.1186/s12884-023-06238-5.

11. LE S, S W, C K, B P, HL. H. Use of Maternal Early Warning Trigger tool reduces maternal morbidity. Obstet and Gynaecol. 2016;(214):6.

12. El Ayadi AM, Nathan HL, Seed PT, Butrick EA, Hezelgrave NL, Shennan AH, et al. Vital Sign Prediction of Adverse Maternal Outcomes in Women with Hypovolemic Shock: The Role of Shock Index. PLoS One. 2016;11(2):e0148729. Epub 20160222. doi: 10.1371/journal.pone.0148729. PubMed PMID: 26901161; PubMed Central PMCID: PMCPMC4762936.

13. Lee SY, Kim HY, Cho GJ, Hong SC, Oh MJ, Kim HJ. Use of the shock index to predict maternal outcomes in women referred for postpartum hemorrhage. Int J Gynaecol Obstet. 2019;144(2):221–4. Epub 20181210. doi: 10.1002/ijgo.12714. PubMed PMID: 30447073.

14. Kruk ME, Leslie HH, Verguet S, Mbaruku GM, Adanu RMK, Langer A. Quality of basic maternal care functions in health facilities of five African countries: an analysis of national health system surveys. The Lancet Global Health. 2016;4(11):e845–e55. doi: 10.1016/S2214-109X(16)30180-2.

15. Andrikopoulou M, D’Alton ME, editors. Postpartum hemorrhage: early identification challenges. Seminars in perinatology; 2019: Elsevier.

16. Beňová L, Semaan A, Portela A, Bonet M, van den Akker T, Pembe AB, et al. Facilitators and barriers of implementation of routine postnatal care guidelines for women: A systematic scoping review using critical interpretive synthesis. J Glob Health. 2023;13:04176. Epub 20231124. doi: 10.7189/jogh.13.04176. PubMed PMID: 37997894; PubMed Central PMCID: PMCPMC10668206.

17. Health Mo. Essential Maternal and Newborn Clinical Care Guidelines for Uganda. 2022.

18. Mbale Regional Referral Hospital. Available from: https://mbalehospital.go.ug/.

19. Ministry Of Health U. Essential Maternal Newborn Care Guidelines 2022 2022.

20. Mugyenyi GR, Ngonzi J, Wylie BJ, Haberer JE, Boatin AA. Quality of vital sign monitoring during obstetric hospitalizations at a regional referral and teaching hospital in Uganda: an opportunity for improvement. Pan Afr Med J. 2021;38:252. Epub 20210311. doi: 10.11604/pamj.2021.38.252.21749. PubMed PMID: 34104300; PubMed Central PMCID: PMCPMC8164430.

21. Beňová L, Semaan A, Portela A, Bonet M, van den Akker T, Pembe AB, et al. Facilitators and barriers of implementation of routine postnatal care guidelines for women: A systematic scoping review using critical interpretive synthesis. Journal of global health. 2023;13:04176.

22. Kairithia F, Karanja G. Joseph, Eunice C, Kinuthia J, Chege M, et al. Adequacy of vital signs monitoring in post delivery mothers at the Naivasha District Hospital of Nakuru County, Kenya. International Journal of Medical and Clinical Sciences. 2014.

23. Agaba DC, Lugobe HM, Migisha R, Jjuuko M, Saturday P, Kisombo D, et al. Abnormal obstetric shock index and associated factors among immediate postpartum women following vaginal delivery at a tertiary hospital in southwestern Uganda. BMC Pregnancy Childbirth. 2024;24(1):31. Epub 20240104. doi: 10.1186/s12884-023-06238-5. PubMed PMID: 38178057; PubMed Central PMCID: PMCPMC10768342.

